# Experiences of psycho-oncological counselling to support mental health in South Australians diagnosed with cancer

**DOI:** 10.1101/2023.08.06.23293728

**Authors:** Jason Blunt, Joshua Trigg

**Author notes:** Corresponding author; PO Box 2100, Adelaide SA 5100. **Declaration of competing interest** The authors declare that they have no known competing financial interests or personal relationships that could have appeared to influence the work reported in this paper.

## Abstract

**Background:** Effective delivery of psycho-oncological support requires understanding of client perceptions of counselling service effectiveness, psychosocial outcomes, and meeting of client support needs and expectations.

**Objective:** This study aimed to describe perceptions of clients accessing psycho-oncological counselling for people directly or indirectly affected by cancer, and describe perceived psychological distress, depression, and anxiety from pre-to post-counselling.

**Methods:** South Australian psycho-oncological counselling service clients were recruited (*n*=28). Psychological distress, anxiety, and depression were assessed before and after counselling sessions. Client expectations, experiences, and counselling outcomes were examined via pre-post-tests, and thematic analysis.

**Results:** Clients reported reduced anxiety (*t*=-2.31, *p*=.029), depression (*t*=-2.51, *p*=.018), distress (*t*=-4.19, *p*<.0001), and global mental health symptomology (*t*=-2.79, *p*=.009). Four themes were identified: having no expectations, needing help managing emotions, seeking coping strategies, and seeking better understanding of their experience. Client expectations were satisfied (92.8%), regardless of counselling reason.

**Conclusion:** Benefits of counselling included reduced symptomology, receipt of knowledge and skills, and increased ability to manage everyday life. Supportive counselling significantly reduces distress and symptoms of anxiety and depression while supporting client and family functioning during cancer treatment.

**Implications:** Individual supportive counselling plays an integral role in lives of cancer patients and family members. Clients face concerns relating to cancer prognosis (e.g., recurrence fear), and to broader related experiences (e.g., social dynamics). Complex needs across cancer experience as a patient, carer, or other family member, requires that psycho-oncological counselling targets major client expectations, promotes benefits of counselling, and strategies for managing daily life events.

## 1. Introduction

### 1.1 Purpose/objectives

Despite sustained investment in cancer prevention, education, support and treatment, more than a million Australians are either receiving treatment, or have survived cancer, with 150,000 diagnosed with cancer annually.^1^ Living with cancer presents multiple psychosocial challenges to patients and people in their supporting relationships.^2–5^ Influential psychosocial care considerations during the cancer journey include experiencing fear, anxiety, and depression, fatigue, pain, or cognitive impairment, sexual dysfunction, social isolation, or employment and financial issues.^3^ Among these considerations, mental health features prominently, with a 32% prevalence of psychological conditions in cancer patients.^6^

Beyond improving quality of life generally, psychosocial care promotes treatment compliance, reduces or prevents anxiety and depression symptoms, and helps patients manage their diagnosis and its consequences.^6^ Programs designed to support patients managing the psychosocial sequelae of cancer are increasingly available, with counselling-based interventions linked with improved coping, quality of life, and distress alleviation.^7, 8^ Despite being broadly implemented, and current evidence supporting efficacy and satisfaction of psycho-oncological support groups, research focusing on individual psycho-oncological counselling remains scarce, even in the context of patient-oriented medicine.^9–12^ Individual psychological support can be more appropriate for people who prefer one-on-one support, or who have limited capacity (or comfort) to participate in structured group sessions. The benefits of individual therapy, compared to support groups, are largely unreported from perspectives of cancer patients, highlighting a key knowledge gap in psycho-oncological care.^9^

Cancer patients emphasise that as the diagnosis restructures their life, they expect counselling to help with managing new problematic situations, and their capacity to express support needs to professional caregivers.^7^ In one case of a crisis and supportive counselling service, focusing on these needs helped meet support expectations for 84% of clients.^7^ Similarly, evaluation of individual outpatient psycho-oncological counselling identified four factors that shape patients’ experience: distress management, service access challenges, service benefits, and the therapeutic encounters.^8^ However, by focusing on early counselling phases only^8^, rather than the completed experience,^7^ we are offered an incomplete picture of client perceptions of psycho-oncological support services.

Pairing qualitative and quantitative description of client support experiences further builds this picture through measuring mental health changes associated with service access. Studies show the need to record baseline mental health indicators of distress, anxiety, and depression, though can neglect to re-assess post-intervention indicators.^13^ The crucial role of psycho-oncological counselling is supported by a large randomised multisite trial for cancer patients with major depression. Patients who received individual psycho-oncological counselling in conjunction with oncological care reported significantly improved mental health and quality of life, with eight-times greater the likelihood (AOR=8.5, 95%CI5.5-13.4) of halved depression symptoms at 24 weeks if receiving counselling.^14^ At 12-months post-counselling, this included lower depression, anxiety, pain, and fatigue, and significantly improved quality of life.^14^ As cancer patients present with symptoms ranging beyond depression, counselling services need to encompass a variety of causes and levels of distress.

### 1.2 Psycho-oncological counselling service

The Psycho-oncological Counselling Service (PCS) available via Cancer Council South Australia supports people directly or indirectly affected by cancer, including patients, partners, carers, family, friends, and colleagues. It is accessed by calling an information line (13 11 20) and requesting or being referred to the PCS. Counselling is offered at any cancer stage, at diagnosis, during and after treatment, to advanced cancer care, bereavement, and survivorship. One-hour counselling sessions are provided free of charge in-person or as telehealth sessions (e.g., telephone, videocall).

Confidential counselling sessions provide support strategies in multiple domains: 1) managing negative thoughts; 2) understanding and expressing emotions helpfully; 3) identifying and clarifying necessary decisions and choices; 4) communicating with family and friends about concerns; 5) finding new ways to manage stress, set goals and achievement strategies; and 6) discussing realistic hope support in managing grief. The PCS is staffed by qualified counsellors eligible for Australian Psychological Society, Australian Association of Social Workers, or Psychotherapy and Counselling Federation of Australia membership. Given the important role of the PCS in South Australia’s psycho-oncological care ecosystem, its regular evaluation is essential to ensuring that high-quality supportive counselling is available.

### 1.3 Aims

This study aimed to describe perceptions of clients accessing community psycho-oncological counselling designed for people directly or indirectly affected by cancer, and to describe changes in clients perceived psychological distress, depression, and anxiety from pre-to post-counselling. It was anticipated that counselling supports positive mental health, and that changes in these indicators would be seen. The study also classified PCS users to identify which aspects of the service contributed to its effectiveness.

## 2 Methods

### 2.1 Participants

Study involvement was open to all clients referred to the counselling service between 01/12/2018 and 31/03/2020. Overall, 115 participants were recruited prior to their first appointment, of which 28 participants (82% female) completed post-counselling follow-up measures. Attrition is noted in the discussion, though responders and non-responders were similar in gender, client type and cancer type, and the number of counselling sessions attended. However, non-responders were significantly younger and reported significantly higher initial distress, compared to the responder group (Table 1).

Prior to counselling, clients were administered the Distress Thermometer, and the Hospital Anxiety and Depression Scale (HADS) by the counsellor. The distress thermometer, a single item that screens patients for distress level, rated from 0 (no distress) to 10 (extreme distress), has been extensively used with oncology patient populations.^15, 16^ The HADS, is a 14-item self-report screening tool for assessing emotional distress, state anxiety and depression in clinical and non-clinical populations, with sound psychometric properties in detecting anxiety and depression.^17, 18^ Items are scored on a four-point scale with total scores per subscale ranging from zero to twenty-one, with higher scores indicating higher levels of anxious of depressive state. After their final session, participants received a mailed paper copy or emailed link to the follow-up survey measures including the distress thermometer and HADS, and qualitative prompts from previous research.^7^ Developed to describe cancer patients’ perceived outcomes of a counselling service, this 25-item 15- minute survey focused on prior expectations, perceived benefits, and outcomes achieved by attending the service^7^ (Appendix A).

### 2.2 Procedure

Research ethics approval was obtained from the Cancer Council Victoria Human Research Ethics Committee (CCVIC HREC: 1808). On first contacting Cancer Council SA Information Service, and having study involvement described to them, participants completed separate verbal informed consent for participation prior to referral to the PCS. The study process was modelled on a Cancer Council SA evaluation. Information Service nurses informed clients of the ongoing study and the process for collecting information via a script. A distress thermometer reading was also recorded, to control for the natural variation in symptomatic distress between initial call with the nurses and the first counselling appointment.

Demographic information, recorded by nurses to a client database, was extracted for those clients who consented to participate. Data were collected in this way to minimise participant and counsellor burden. At their first counselling session, participants were reminded of the study, and the distress thermometer and HADS were administered.

Following the final counselling session (as identified by their counsellors), participants completed post-counselling measures either in the room once the counsellor had departed, or by return mail to the researchers. The 25-item post-counselling survey took 15 minutes to complete

### 2.5 Data analysis

Descriptive data analysis included the use of frequencies, chi-square, and paired sample *t*- tests. Analyses were conducted only on cases with complete data for target variables, and this was done using SPSS v.23. Thematic analysis of qualitative data was conducted using a realist approach to report participant experiences and meanings as described. This followed open and axial coding of brief open-ended responses using a custom Excel worksheet.^19^ Qualitative analysis followed Consolidated Criteria for Reporting Qualitative research guidelines.

## 3 Findings

### 3.1 Participant demographics

Table 1 shows participants were mostly female, averaged 50-60 years of age, and predominately cancer patients (responders=67.9%, non-responders=51.7%) or family members (responders=17.9%, non-responders=27.6%). Participants averaged over 2 counselling sessions, primarily relating to breast (responders=35.7%, non-responders=19.5%%) or bowel (responders=10.7%, non-responders=17.2%) cancer types.

### 3.2 Mental health indicators

Changes in state distress, depression, anxiety, and overall mental health impairment were assessed via HADS scores. Participants reported significantly lower general distress following receipt of counselling sessions through the counselling service, a result which was not affected by the number of sessions completed: 1 session (M±SD=4.9±3.1), 2-3 sessions (M±SD=2.3±2.1), or 4+ (M±SD=3.1±2.3) (*F*(2, 25) = 2.45, *p* = .109).

**Table 1.**
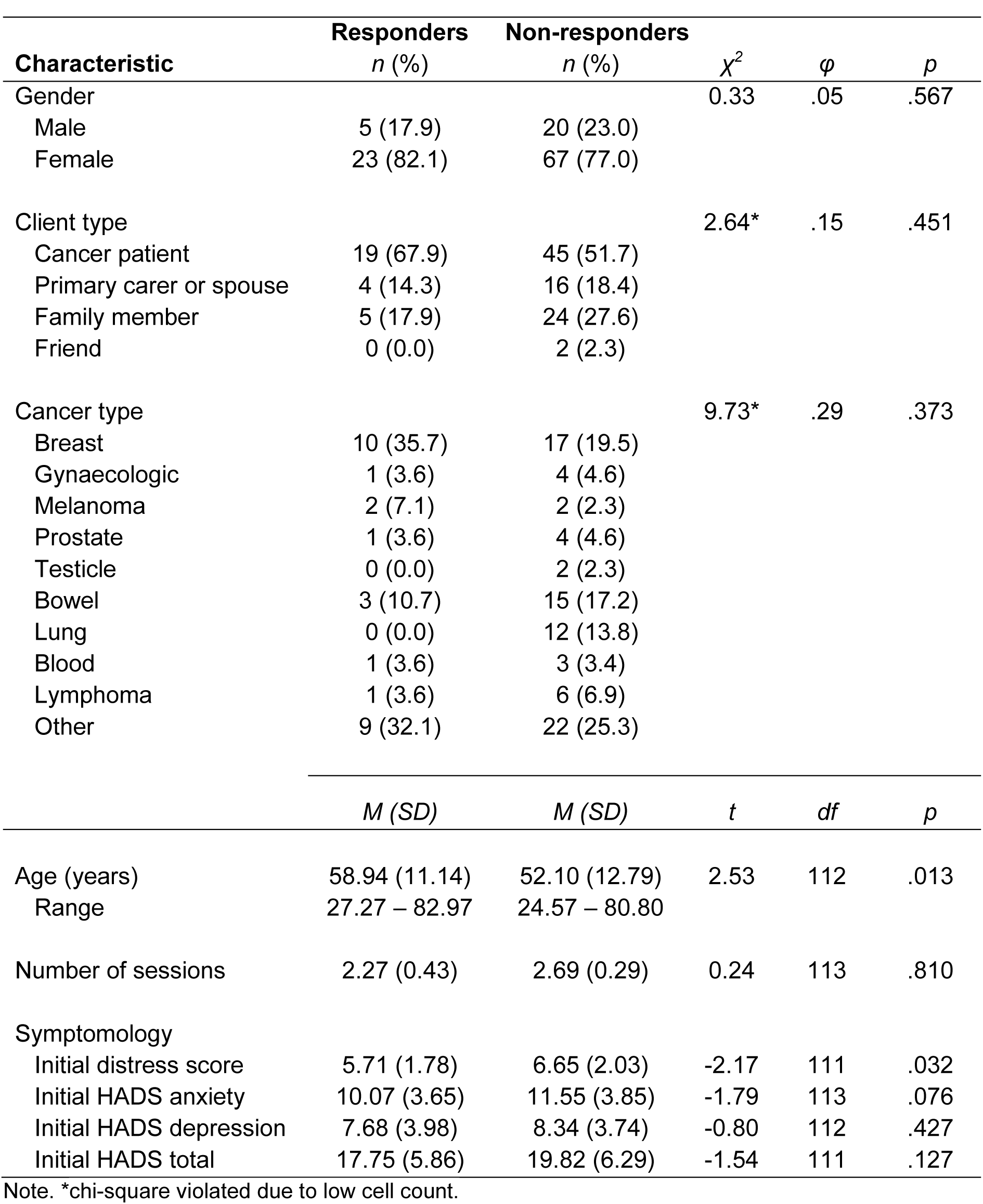
Demographics of counselling service study responders (n=28) and non-responders (*n*=87).

| Characteristic | Responders<br>*n* (%) | Non-responders<br>*n* (%) | $\chi^2$ | $\phi$ | *p* |
| --- | --- | --- | --- | --- | --- |
| Gender |  |  | 0.33 | .05 | .567 |
| Male | 5 (17.9) | 20 (23.0) |  |  |  |
| Female | 23 (82.1) | 67 (77.0) |  |  |  |
| Client type |  |  | 2.64* | .15 | .451 |
| Cancer patient | 19 (67.9) | 45 (51.7) |  |  |  |
| Primary carer or spouse | 4 (14.3) | 16 (18.4) |  |  |  |
| Family member | 5 (17.9) | 24 (27.6) |  |  |  |
| Friend | 0 (0.0) | 2 (2.3) |  |  |  |
| Cancer type |  |  | 9.73* | .29 | .373 |
| Breast | 10 (35.7) | 17 (19.5) |  |  |  |
| Gynaecologic | 1 (3.6) | 4 (4.6) |  |  |  |
| Melanoma | 2 (7.1) | 2 (2.3) |  |  |  |
| Prostate | 1 (3.6) | 4 (4.6) |  |  |  |
| Testicle | 0 (0.0) | 2 (2.3) |  |  |  |
| Bowel | 3 (10.7) | 15 (17.2) |  |  |  |
| Lung | 0 (0.0) | 12 (13.8) |  |  |  |
| Blood | 1 (3.6) | 3 (3.4) |  |  |  |
| Lymphoma | 1 (3.6) | 6 (6.9) |  |  |  |
| Other | 9 (32.1) | 22 (25.3) |  |  |  |
|  | *M* (*SD*) | *M* (*SD*) | *t* | *df* | *p* |
| Age (years) | 58.94 (11.14) | 52.10 (12.79) | 2.53 | 112 | .013 |
| Range | 27.27 – 82.97 | 24.57 – 80.80 |  |  |  |
| Number of sessions | 2.27 (0.43) | 2.69 (0.29) | 0.24 | 113 | .810 |
| Symptomology |  |  |  |  |  |
| Initial distress score | 5.71 (1.78) | 6.65 (2.03) | -2.17 | 111 | .032 |
| Initial HADS anxiety | 10.07 (3.65) | 11.55 (3.85) | -1.79 | 113 | .076 |
| Initial HADS depression | 7.68 (3.98) | 8.34 (3.74) | -0.80 | 112 | .427 |
| Initial HADS total | 17.75 (5.86) | 19.82 (6.29) | -1.54 | 111 | .127 |

Note. \*chi-square violated due to low cell count.

**Table 2.**
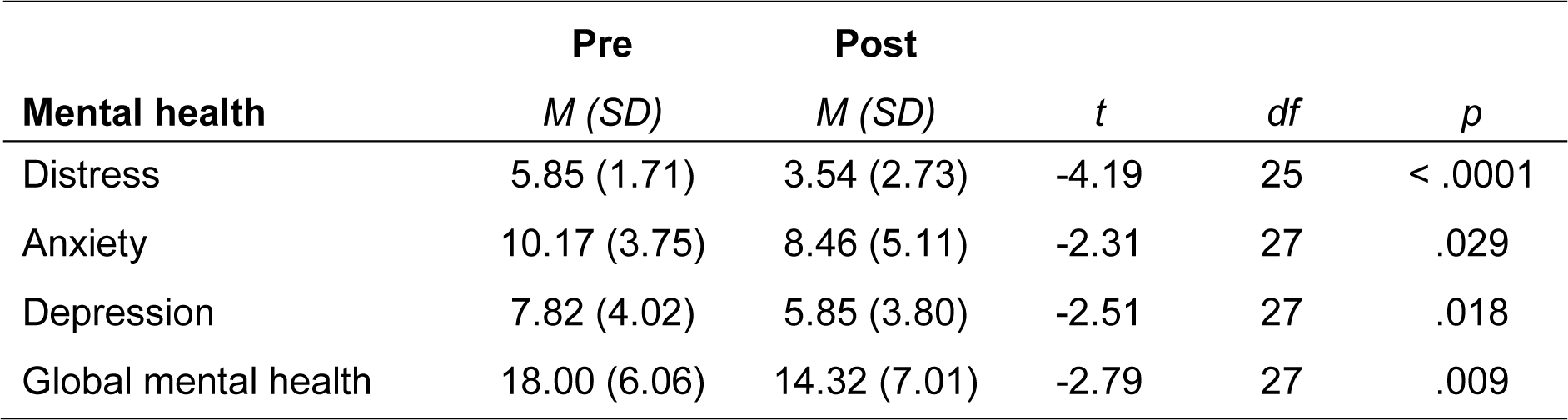
Pre- and post-counselling measures of distress, depression, and anxiety for responders (*n*=28).

| **Mental health** | **Pre** | **Post** | *t* | *df* | *p* |
| --- | --- | --- | --- | --- | --- |
|  | *M (SD)* | *M (SD)* |  |  |  |
| Distress | 5.85 (1.71) | 3.54 (2.73) | -4.19 | 25 | < .0001 |
| Anxiety | 10.17 (3.75) | 8.46 (5.11) | -2.31 | 27 | .029 |
| Depression | 7.82 (4.02) | 5.85 (3.80) | -2.51 | 27 | .018 |
| Global mental health | 18.00 (6.06) | 14.32 (7.01) | -2.79 | 27 | .009 |

### 3.3 Supportive conversations

Participants were asked if they had intimate supportive conversations with other people concerning their or their family member’s cancer illness. Of the 23 (82.1%) who responded that they had, four (14.3%) indicated they spoke to one to two people, seven (25%) indicated three to four, five (17.8%) spoke to four or five people, and four (14.4%) indicated eight or more. Most indicated they had spoken with family and close friends (*n* = 20, 71.4%), health staff (*n* = 11, 9.3%), friends or acquaintances (*n* = 6, 21.4%), workmates (*n* = 4, 14.2%), fellow patients (*n* = 3, 10.7%), and others (*n* = 2, 7.1%).

### 3.4 Client expectations

Clients were reported their service expectations prior to counselling. Four key themes were identified in clients’ expectations of the counselling service: *no expectations*, *help dealing with feelings and emotions*, *specific strategies for coping*, and *better understanding*. Clients often expressed practical expectations regarding engaging with the service. Responses focused on the need for strategies for dealing with mental health issues, such as alleviating depression, managing cancer support, processing diagnoses and related stressors (e.g., medication side effects). Seeking help to deal with emotions and feelings was also a common reason for accessing the service, including processing feelings about diagnoses, to expressing feelings in a safe and supportive therapeutic environment:

> *I wanted to be able to say things out loud that I could not share with my family about fear and death*.
>
> *To discuss my concerns with a person with knowledge of cancer issues but who had no vested emotional interest in me*.
>
> *A listening ear. An opportunity to explore my feelings regarding my situation*.
>
> *To being supported in a safe environment and freely discuss what I was feeling*.

Clients with no expectations included those who simply reported no expectations, and those whose expectations were limited by their uncertainty around how effective counselling would be for them. This centred-on time constraints, and doubting counselling effectiveness:

> *I didn’t think it would be helpful as I was too busy working…*
>
> *I really did not know what to expect going into it, but felt it was important to try it out*.
>
> *I was unsure how helpful it would be. When my son was first diagnosed, I paid for professional counselling, and was asked many questions which I did not think were relevant*.

These expectations of the counselling service were fulfilled for most clients (*n* = 26, 92.8%). When asked how satisfied they were with their experience with the counselling service, 12 (42.8%) were *more than satisfied*,12 (42.8%) were *satisfied*, two (7.6%) were *rather satisfied*, with two (7.6%) responding they were *disappointed*. Satisfaction with the counselling experience related to the counsellor and the comfort realised through the therapeutic alliance:

> *She was a good listener, patient, helpful and supportive*.
>
> *The therapist that I met with was lovely and supportive and empathetic and understanding*.
>
> *She helped me to understand that it would help my son and his daughter, by looking after my own wellbeing…. I am grateful for her experience, her warm accepting manner, listening, helpful understanding of families*.

Clients expressed themes of comfort, particularly regarding normalisation through the commonality of experiences shared amongst cancer sufferers:

> *Always made me feel secure and good about myself and that I was progressing even though it was slow, it was steady, and that I would win in time*.
>
> *…basically, I felt some of my feelings were normalised, and that helped*.

Techniques and strategies for coping were also particularly satisfying aspects of the counselling process:

> *I learned some very useful and unexpected techniques to help me cope with life in general, cancer in particular*.
>
> *Good advice with some practical suggestions to meet my needs at different stages*.
>
> *Counselling really helped me reset and activate some helpful things to do, e.g., acupuncture, general self-care, and asking for help*.

Two clients who expressed that their expectations were not met did not comment further.

### 3.5 Benefits of counselling

Participants indicated how they had benefited from counselling using a predefined list of commonly expected outcomes derived from previous literature.^7^ The most reported benefits were *opportunity to practice talking about my life situation* (*n* = 21, 77.8%) and an *increased understanding of their reactions and feelings* which they experienced during the process of their (or their family member’s) cancer illness (*n* = 20, 74.1%). Feeling that the *counsellor saw them as a person* (*n* = 18, 66.7%), *feeling more initiative* (*n* = 11, 40.7%), and *feeling less fear* (*n* = 10, 37%) were also commonly reported benefits. Several participants indicated feeling an *increased distance* from their situation (*n* = 5, 18.6%), *appreciating the joys of everyday life* from a more profound perspective (*n* = 5,18.6%), and becoming less fatigued (*n* = 5, 18.6%) after participating in counselling. Other benefits (*n* = 2, 7.1%) expressed by clients included feeling *more prepared for the reality of cancer* and *feeling lighter*.

### 3.6 Managing everyday situations

Clients were asked whether the counselling helped them to manage typical daily life situations. Most respondents indicated that it did (*n* = 21, 77.8%), with 11 (57.9%) indicated it was *much better*, and eight (42.1%) indicating *somewhat better* (9 participants gave no response). When asked for a clear example of a situation in which counselling had benefited them, clients described situations relating to three themes: *coming to terms with their situation*, *grieving loved ones*, and the *use of psychological skills* taught to them by the counsellors. Coming to terms reflected clients taking their diagnosis one step at time, gaining comfort with their diagnosis, not delaying treatment to wait for new approaches, and the emotional release associated with counselling:

> *Prior to counselling I often became overwhelmed, teary, and emotional as I thought of my circumstance. Being able to discuss the issues seems to have given me the emotional release I needed*.

Another commonly mentioned theme, grieving loved ones, was based on statements made around being able to talk about death and loss openly, fear of losing a spouse/partner, being able to think about lost loved ones, and being able to view aspects of the past differently following counselling sessions:

> *I was able to talk about death in the open and the chance I may become a widow earlier than I ever imagined. After I said it out loud it stopped going around in my head as much - the fear was less*.

### 3.7 Other beneficial activities

Clients were asked if there was anything else they had done since their first counselling session which had helped them. Two main themes were identified: *accessing further services*, and *increased spirituality*. Further service access was common, particularly seeking further physical and mental therapy for cancer-specific side effects, such as the physical impacts of chemotherapy, and further cancer-specific anxiety treatment. *Spiritualty* was reflected in increased practises or renewal of traditional faiths, and adoption of alternative modalities such as mindfulness practice, ‘talking’ to lost loved ones, naturopathy, and massage. Other comments regarding the counselling experience emphasised gratitude for the counsellors and the free support they provide.

## 4 Discussion

Client perceived outcomes and mental health status were assessed following use of a psycho-oncological counselling service (PCS). Pre-and post-service utilisation measures assessed change in participant levels of distress, anxiety, depression, and global mental health. This study also collected fixed-choice and open-ended qualitative feedback to describe client experiences and satisfaction with the PCS

Distress, depression, and anxiety symptoms were significantly reduced following the receipt of psycho-oncological counselling. The decrease in global mental health symptomology highlights the beneficial impact that short counselling interventions can have on clients’ psychological wellbeing.^20^ Clients reported significantly lower general distress following counselling sessions, a result which was not affected by the number of sessions attended. These findings suggest that individual psycho-oncological counselling, like group therapies, can support the psychological resilience in people affected by cancer.^7, 8, 13^

Clients show a range of expectations when presenting to this type of PCS, ranging from no expectations, help in dealing with feelings and emotions, to specific coping strategies and better understanding of their experience. Despite mixed experiences, expectations of the psycho-oncological counselling service were satisfied in most cases. Reduced symptomology, receipt of beneficial knowledge, and increased ability to manage their everyday lives, supported by client satisfaction with the service, highlight the unique and important role of a supportive counselling service to cancer patients and their families. Given this, future research or evaluations can examine expectations outside of these core themes to identify new areas for support that can support client psycho-oncological resilience.

The major themes of our analysis align with previous work to indicate that the major outcome of counselling was assistance to work through the crisis and to find meaning or an authentic voice in the suffering involved with cancer.^7^ Being able to manage problems and reformulate the situation in relation to the illness are important and established outcomes in counselling interventions,^7^ seen in this study via reduced adjustment-related symptomology. PCS clients strongly indicated that multidomain support benefits can be obtained from a small number of sessions, which may in turn confer benefits to cancer patients’ and supports’ wider social network (e.g., family members). Most PCS clients viewed the counsellors highly positively, with appreciation for professional and individually tailored care approaches reflected in additional client feedback.

Although all clients accessing the PCS being invited to participate, the low response rate, (24%), though reflective of oncological survey research,^21^ was a limitation. Significant differences were evident between responders and non-responders, with responders likely to be older and have lower initial distress than non-responders. This may suggest a population with poorer mental health symptoms that are not being adequately engaged via psycho-oncological counselling. Discussions with the counselling team during the follow-up period indicated that clients with higher distress were often palliative clients or their family members, which in turn made follow-up difficult due to either the client passing, or the inappropriateness of contacting a spouse or family member to collect data. Older clients’ higher responsivity has been noted in previous psycho-oncological counselling research.^7, 20^ Yet, it remains unclear whether there was something particular about this group (e.g., loss of peer or family support as age increases), or of their younger counterparts (e.g., more accessible peer support, young children/families to attend to), that influenced the likelihood of their seeking support for their cancer journey.

Despite limitations, using a mixed-methods approach that coupled mental health indicator scores with qualitative description of support experiences was a benefit. This design offered unique findings, for a hard to access group, providing insight into PCS client experiences. Given the limitations, this approach enables tentative conclusions to be drawn between key themes of client’s experience of supportive counselling and the drivers of both satisfaction and psychological improvement upon attending the service.

## 5. Implications for psychosocial providers or policy

Practical implications include reinforcing that individual supportive counselling plays an integral role in lives of cancer patients and family members. Clients are often faced with a myriad of concerns, relating to a cancer prognosis (e.g., diagnosis, treatment, surgery, recurrence fear), and to broader related experiences (e.g., body image, sexual functioning, social and family dynamics). This complex interplay of needs across the client’s cancer experience, as a patient, carer, or other family member, requires that psycho-oncological counselling services offer support that targets the major client expectations, promotes the benefits of counselling, and strategies for managing daily life events. Importantly, larger scales intervention and follow-up is needed to confirm these initial findings of reduced distress, anxiety, and depression, and to publicly establish the benefits of psycho-oncological counselling for people affected by a cancer diagnosis.

## Data Availability

All data produced in the present study are available upon reasonable request to the authors.

## Acknowledgments

The authors would like to thank the counselling service clients who opted to take part in this research, as well as the counselling service staff who assisted with data collection.

## Appendix A

### Community Counselling Service Study 2020

We are collecting information from people who use the Cancer Counselling Service in order to ensure that we are providing the best possible service to people who have been affected by cancer. Participation is entirely voluntary and involves completing two brief measures, one which you have already completed and one now that you have completed your final counselling session. If you would like to know more about how this information is used, your counsellor can provide you with further information, or you can contact the Project Officer

If you do choose to participate, completing this questionnaire will only take a few minutes, and any information you provide us is strictly confidential. You do not have to participate if you do not want to, and you are free to withdraw at any time. Your participation or non-participation in this evaluation will in no way impact the service you receive from the counsellors at this service.

On the thermometer diagram to the right, please circle the number (0-10) that best describes how much distress you have been experiencing in the past week including today.

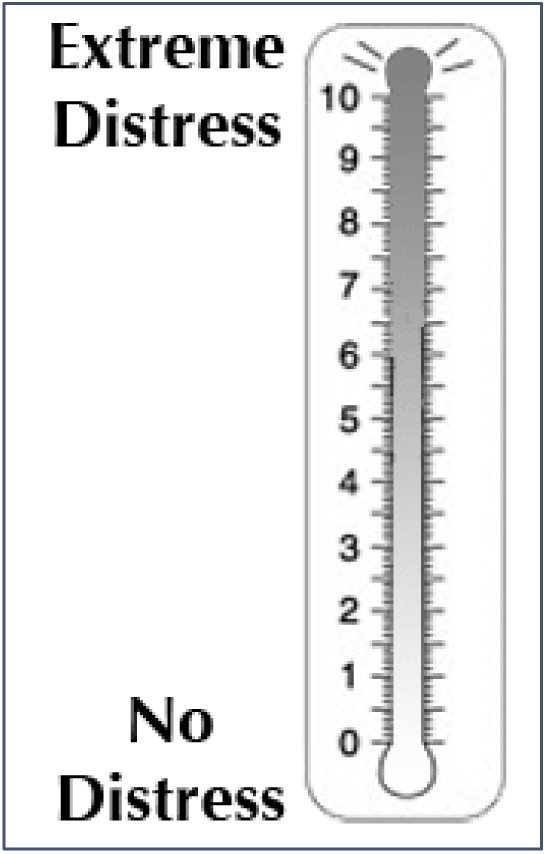

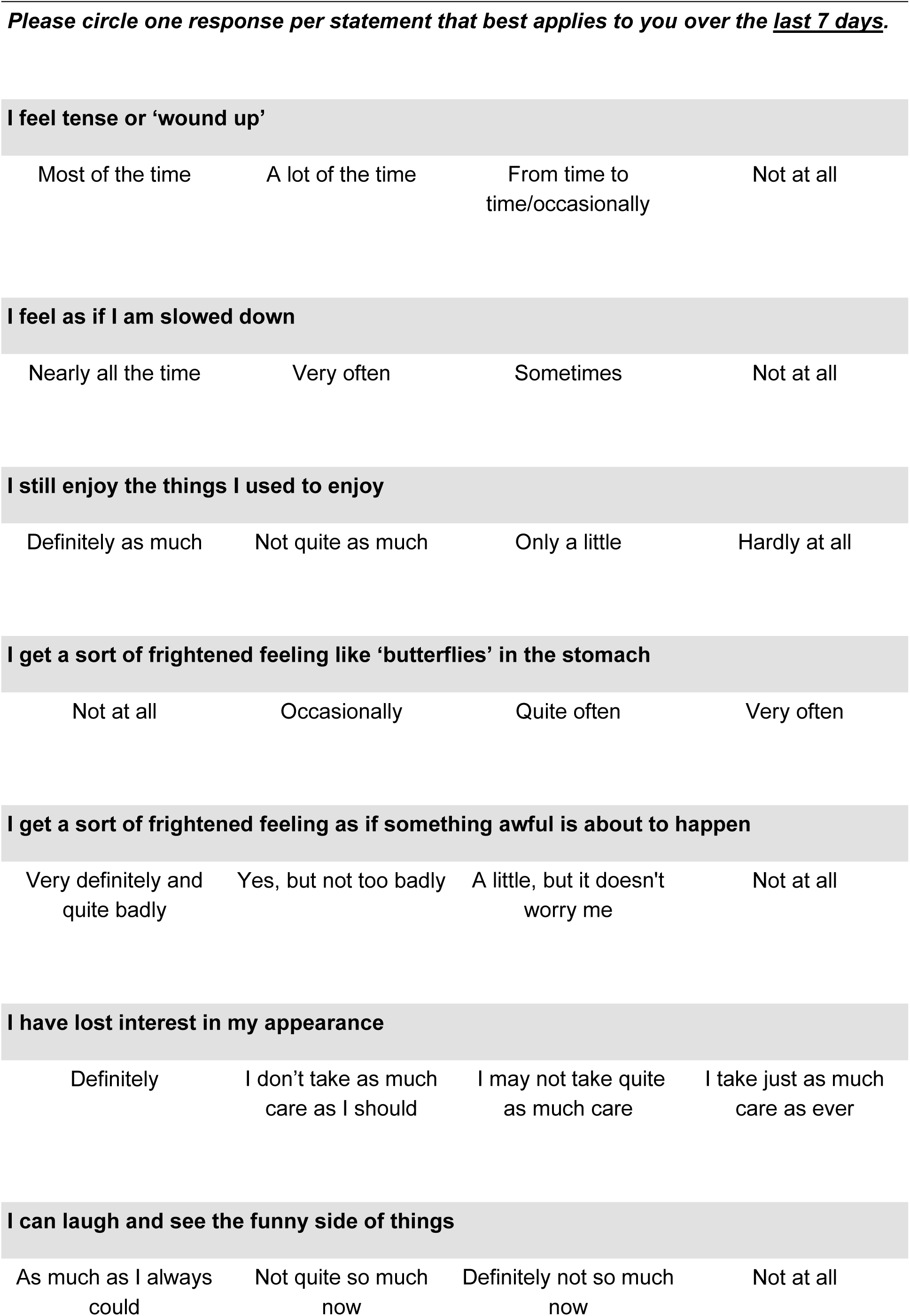
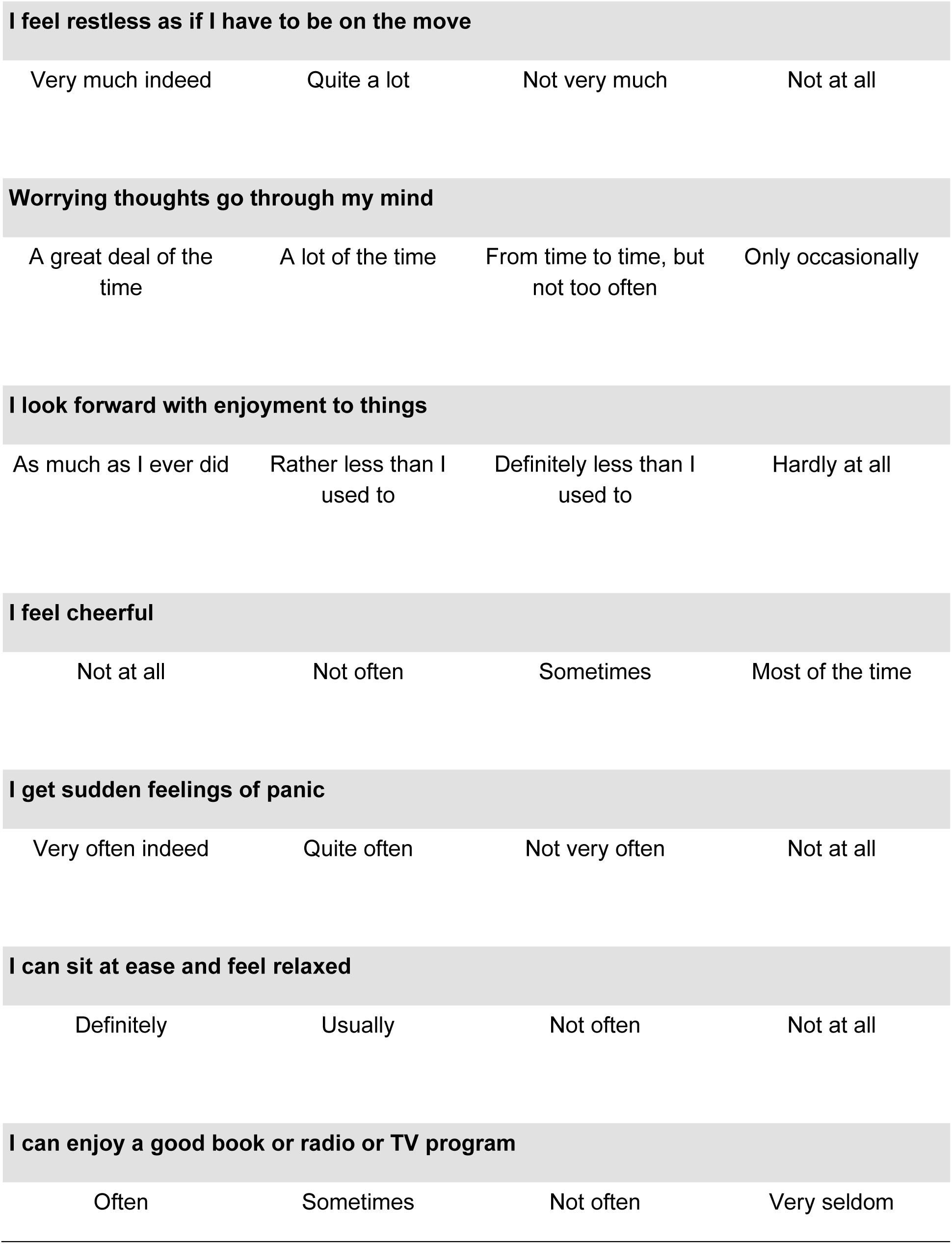
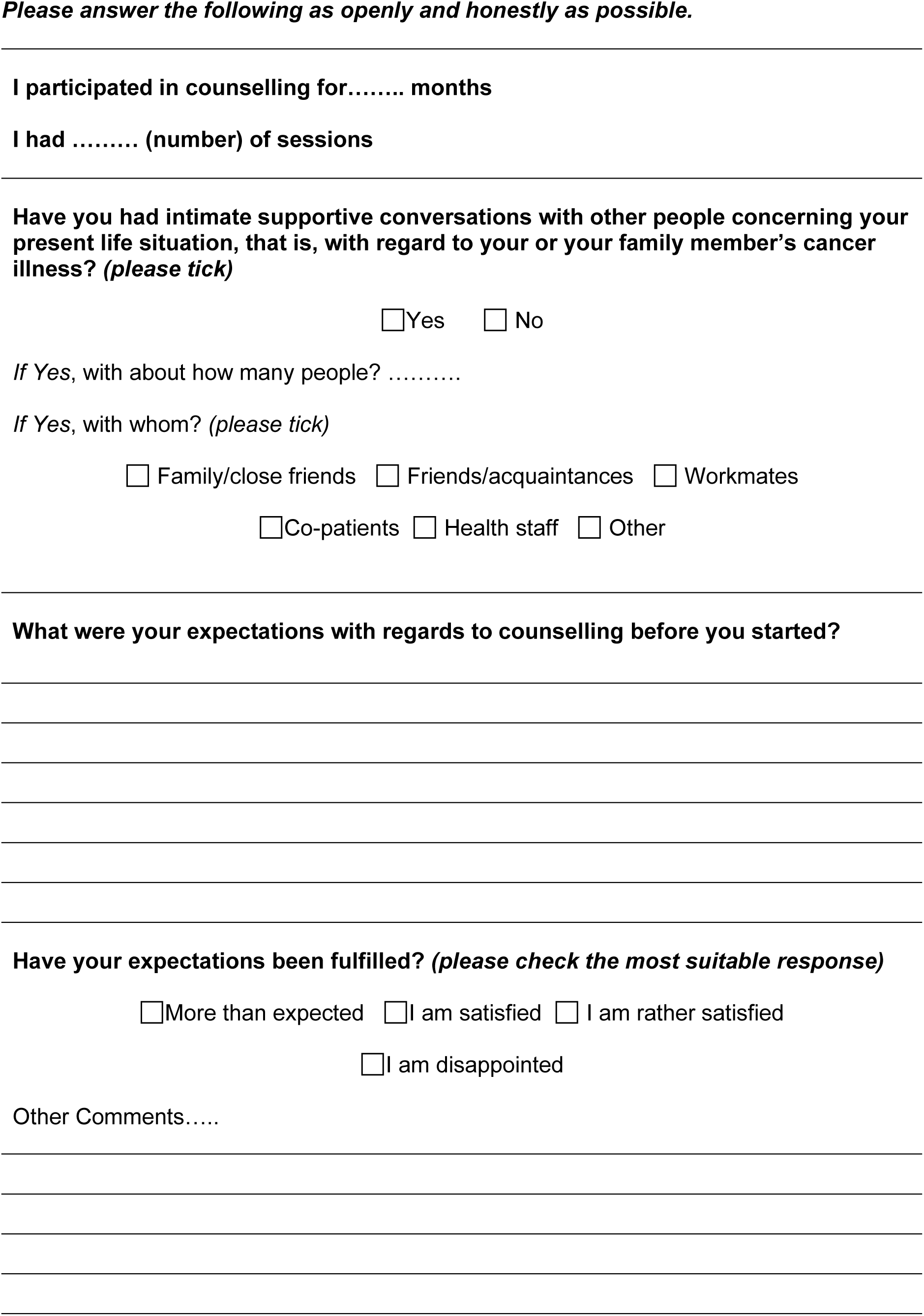
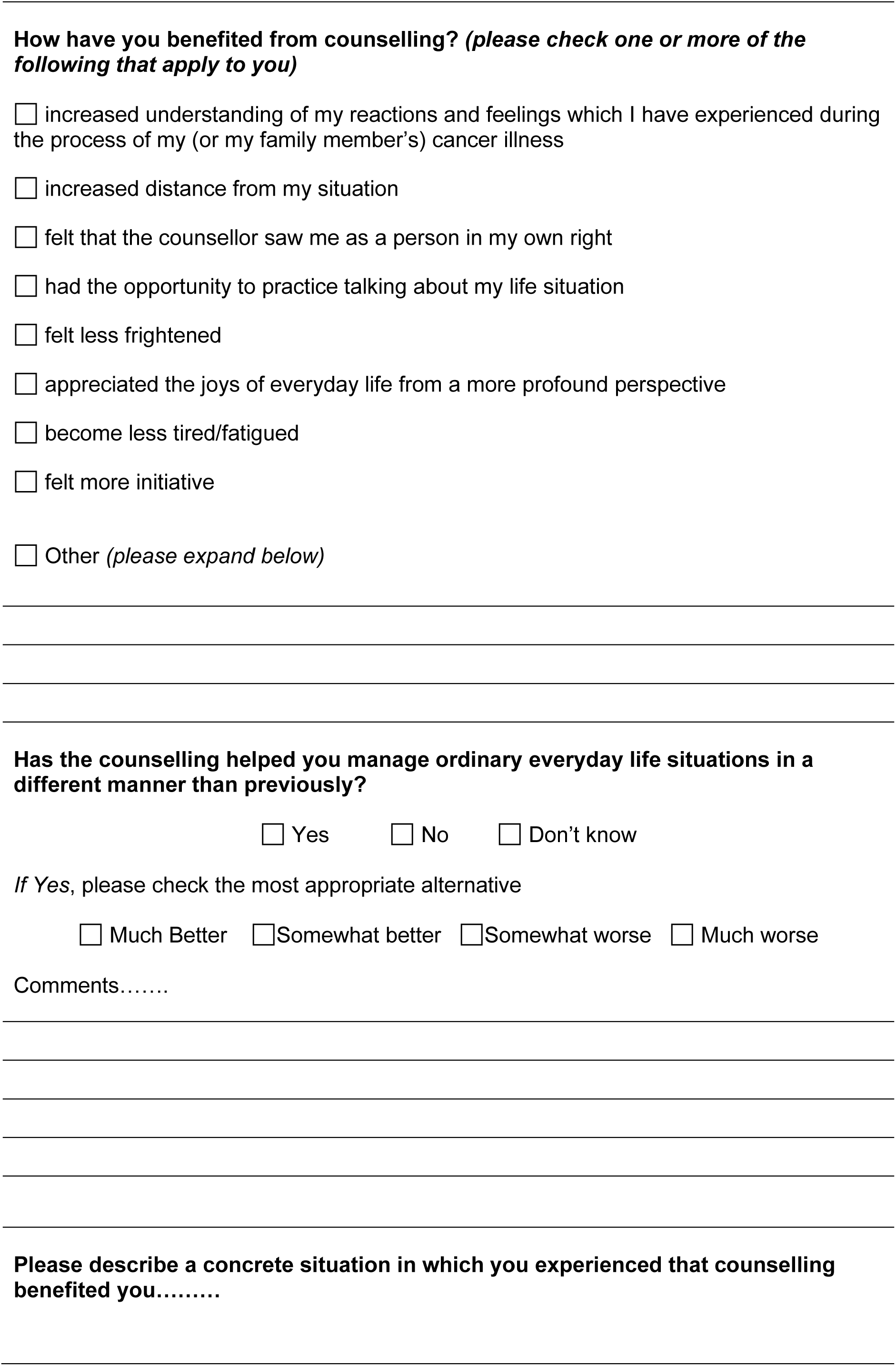
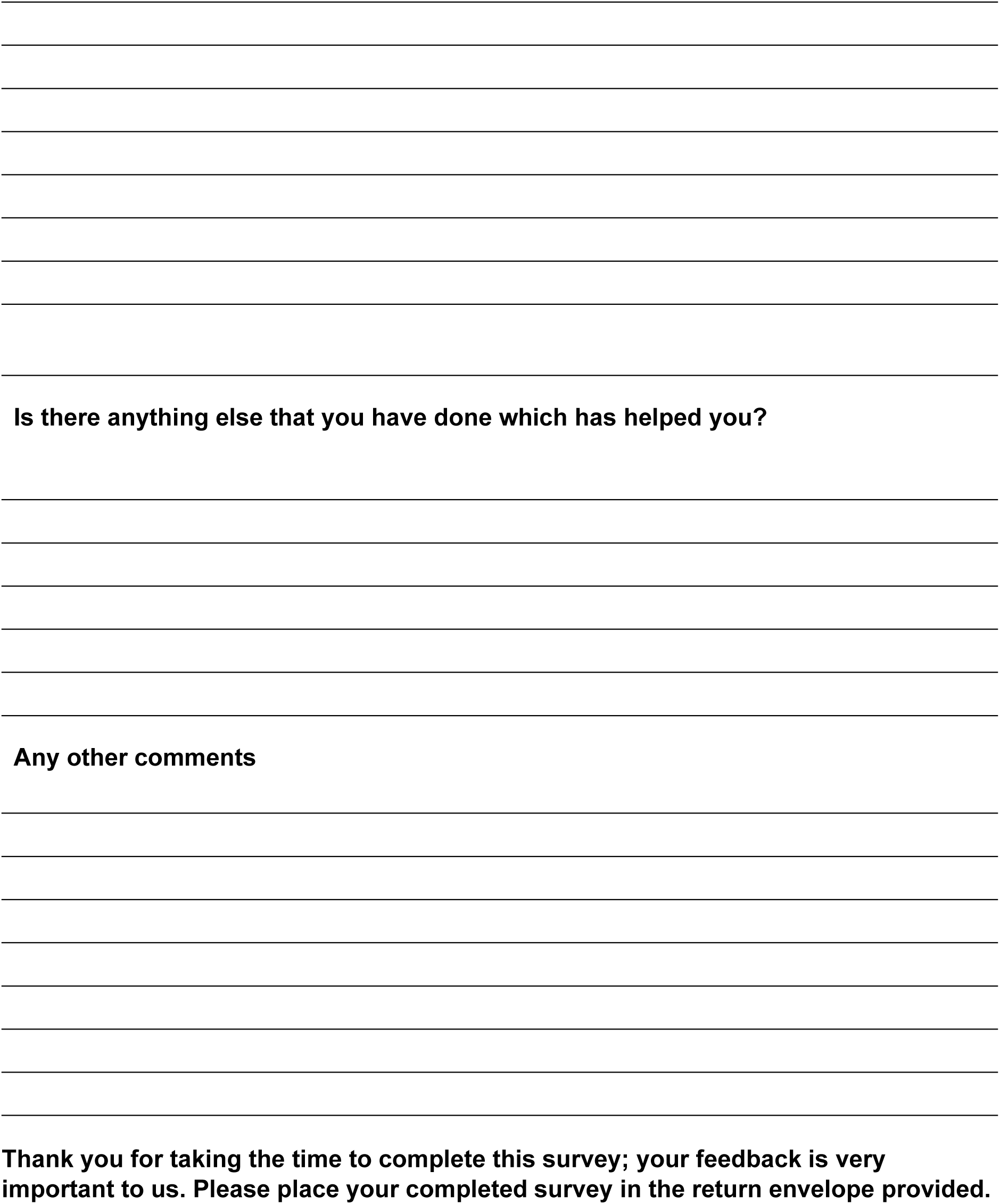

## References

1. Australian Institute of Health and Welfare. Cancer in Australia 2019. Cancer series no. 119. Cat no. CAN 123. Canberra: AIHW;2019.

2. Kowalski C, Ferencz J, Singer S, Weis I, Wesselmann S. Frequency of psycho-oncologic and social service counseling in cancer centers relative to center site and hospital characteristics: Findings from 879 center sites in Germany, Austria, Switzerland, and Italy. Cancer. 2016;122(22):3538–3545.

3. Jacobs LA, Shulman LN. Follow-up care of cancer survivors: challenges and solutions. The Lancet Oncology. 2017;18(1):e19–e29.

4. McGeechan GJ, McPherson KE, Roberts K. An interpretative phenomenological analysis of the experience of living with colorectal cancer as a chronic illness. Journal of clinical nursing. 2018;27(15-16):3148–3156.

5. Singer S, Das-Munshi J, Brähler E. Prevalence of mental health conditions in cancer patients in acute care—a meta-analysis. Annals of oncology. 2010;21(5):925–930.

6. Senf B, Fettel J, Demmerle C, Maiwurm P. Physicians’ attitudes towards psycho-oncology, perceived barriers, and psychosocial competencies: Indicators of successful implementation of adjunctive psycho-oncological care? Psycho-oncology. 2019;28(2):415–422.

7. Öhlén J, Holm A-K, Karlsson B, Ahlberg K. Evaluation of a counselling service in psychosocial cancer care. European Journal of Oncology Nursing. 2005;9(1):64–73.

8. Nekolaichuk CL, Turner J, Collie K, Cumming C, Stevenson A. Cancer patients’ experiences of the early phase of individual counseling in an outpatient psycho-oncology setting. Qualitative health research. 2013;23(5):592–604.

9. Baider L, Peretz T, Hadani PE, Koch U. Psychological intervention in cancer patients: a randomized study. General Hospital Psychiatry. 2001;23(5):272–277.

10. Osborn RL, Demoncada AC, Feuerstein M. Psychosocial interventions for depression, anxiety, and quality of life in cancer survivors: meta-analyses. The International Journal of Psychiatry in Medicine. 2006;36(1):13–34.

11. Sinaiko AD, Landrum MB, Meyers DJ, et al. Synthesis of research on patient-centered medical homes brings systematic differences into relief. Health Affairs. 2017;36(3):500–508.

12. Vaganian L, Bussmann S, Gerlach AL, Kusch M, Labouvie H, Cwik JC. Critical consideration of assessment methods for clinically significant changes of mental distress after psycho-oncological interventions. International journal of methods in psychiatric research. 2020;29(2):e1821.

13. Nekolaichuk CL, Cumming C, Turner J, Yushchyshyn A, Sela R. Referral patterns and psychosocial distress in cancer patients accessing a psycho-oncology counseling service. Psycho-Oncology. 2011;20(3):326–332.

14. Sharpe M, Walker J, Hansen CH, et al. Integrated collaborative care for comorbid major depression in patients with cancer (SMaRT Oncology-2): a multicentre randomised controlled effectiveness trial. The Lancet. 2014;384(9948):1099–1108.

15. O’Donnell E, D’Alton P, O’Malley C, Gill F, Canny Á. The distress thermometer: a rapid and effective tool for the oncology social worker. International journal of health care quality assurance. 2013.

16. Ownby KK. Use of the distress thermometer in clinical practice. Journal of the advanced practitioner in oncology. 2019;10(2):175.

17. Vodermaier A, Linden W, Siu C. Screening for emotional distress in cancer patients: a systematic review of assessment instruments. Journal of the National Cancer Institute. 2009;101(21):1464–1488.

18. Annunziata MA, Muzzatti B, Bidoli E, et al. Hospital Anxiety and Depression Scale (HADS) accuracy in cancer patients. Supportive Care in Cancer. 2019:1–6.

19. Braun V, Clarke V. Using thematic analysis in psychology. Qualitative research in psychology. 2006;3(2):77–101.

20. Boudioni M, Mossman J, Boulton M, Ramirez A, Moynihan C, Leydon G. An evaluation of a cancer counselling service. European Journal of Cancer Care. 2000;9(4):212–220.

21. Parekh AD, Bates JE, Amdur RJ. Response Rate and Nonresponse Bias in Oncology Survey Studies. Am J Clin Oncol. 2020;43(4):229–230.

